# Harnessing multi-output machine learning approach and dynamical observables from network structure to optimize COVID-19 intervention strategies

**DOI:** 10.1101/2024.09.23.24313636

**Authors:** Caroline L. Alves, Katharina Kuhnert, Francisco Aparecido Rodrigues, Michael Moeckel

## Abstract

The COVID-19 pandemic has necessitated the development of accurate models to predict disease dynamics and guide public health interventions. This study leverages the COVASIM agent-based model to simulate 1331 scenarios of COVID-19 transmission across various social settings, focusing on the school, community, and work contact layers. We extracted complex network measures from these simulations and applied deep learning algorithms to predict key epidemiological outcomes, such as infected, severe, and critical cases. Our approach achieved an R2 value exceeding 95%, demonstrating the model’s robust predictive capability. Additionally, we identified optimal intervention strategies using spline interpolation, revealing the critical roles of community and workplace interventions in minimizing the pandemic’s impact. The findings underscore the value of integrating network analytics with deep learning to streamline epidemic modeling, reduce computational costs, and enhance public health decision-making. This research offers a novel framework for effectively managing infectious disease outbreaks through targeted, data-driven interventions.

## I. INTRODUCTION

The COVID-19 epidemic has disclosed difficulties and delays in the public health and societal response to emerging new diseases. While simulation tools designed to model the dynamics of infections have been quickly adapted to new virus parameters [1–3] and made available to a large community of researchers [4], ready-made expert support systems for predicting the effectiveness and impact of public health interventions have not been publicly available. Responsible public health administrators and the general public alike were lacking essential information to opt quickly for the most appropriate and least intrusive intervention.

Hence, a need for new predictive tools has become evident. This paper demonstrates a first direction to further develop agent based disease simulators into tools for predicting optimal public health responses. As a full study of all conceivable interventions is beyond the scope of this work, a focus on contact restrictions is taken.

Mathematical models, notably SEIR (SusceptibleExposed-Infectious-Recovered) models, have been seminal in generating predictions and guiding public health measures [5–7].

Compartmental models are non-linear models widely recognized for their effectiveness in understanding the spread of infectious diseases within a population. The SEIR model subdivides individuals into four distinct compartments based on their disease progression status: Susceptible, Exposed, Infectious, and Recovered [6, 8].

The dynamics of transitions between these compartments are governed by a set of ordinary differential equations (ODEs) driven by essential parameters such as transmission rate, incubation period, and recovery rate described in 1 [8, 9].

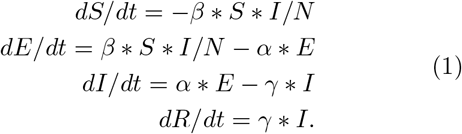

Where: S(t) represents the number of susceptible individuals at time t, E(t) represents the number of exposed individuals (infected but not yet infectious) at time t, I(t) represents the number of infectious individuals at time t, E(t) represents the number of exposed individuals (infected but not yet infectious) at time t, R(t) represents the number of recovered individuals at time t, N is the total population size, *β* is the transmission rate (average number of contacts an infectious person makes per unit time multiplied by the probability of disease transmission in a contact), *α* is the rate at which individuals move from the exposed to infectious compartment (1/incubation period), *γ* is the recovery rate (1/duration of infection).

By leveraging mathematical formulations and incorporating key parameters, compartmental models have proven invaluable in guiding public health measures [5, 10, 11]. On the other hand, agent-based simulators like COVASIM [4] have emerged as powerful tools for studying the complex dynamics of infectious diseases, extending the SEIR model by allowing for the integration of fine grained disease stages, age-specific parameters, immunization models etc, enabling a more nuanced simulation of disease progression across various demographic segments. Furthermore, COVASIM implements the contact network of agents, capturing heterogeneous interaction patterns that the traditional SEIR model oversimplifies, thereby offering a more detailed and realistic representation of disease transmission [12–14].

COVASIM is a stochastic agent-based simulator, which allows for the representation of individual-level heterogeneity in behaviors and interactions, leading to more realistic epidemic predictions [15, 16]. Built on separately generated synthetic populations, this tool additionally includes country-specific demographic information on age structure and population size. Social contact networks are structured in four subgraphs (layers), referring to households, schools, workplaces, and communities [4]. Through COVASIM, it is possible to simulate a refined transmission network by a multigraph with multiple layers, capturing the complex interactions between individuals in a population [13]. The community layer accounts for random contact between individuals in the general population, mimicking casual encounters in public spaces or social gatherings [17]. Household contacts, as another layer, capture close and sustained interactions within households, which are known to be a significant source of disease transmission. The work and school layers further contribute to the complexity of the model as they reflect the specific patterns of interactions that occur in these settings [18]. Workplaces often involve dense and regular contact, while schools accommodate interactions among children and staff, influencing the spread of infections among younger populations [19]. The multigraph contact network enables a detailed and nuanced representation of disease transmission dynamics in various social contexts. The approach allows researchers and policymakers to explore the potential effectiveness of public health interventions in controlling infectious diseases [20–22] in particular those which target specifically the contact networks [13, 23–25].

One major limitation of agent-based simulators like COVASIM is their computational cost [26]. Simulating numerous scenarios with detailed contact networks can require significant computing resources and time.

This work addresses the hypothesis that between a full modeling of contact networks in agent based simulators like COVASIM and neglecting the structure of contact networks in compartment models there is a gap to establish a computationally efficient and fast prediction method which includes some effective properties of contact networks but avoids their full inclusion. We use machine learning for a simplified substitutional model by replacing the full contact network adjacency matrix with a description based on complex network measures, such as betweenness centrality and others based in [27]. They serve as predictors of disease dynamics while other parameters of the COVASIM framework remain fixed.

We use COVASIM to set up the contact network adjacency matrix using its initial state initialization routine, extracted it and calculated relevant complex network measures from it. Those served as input features for a deep learning-based method to predict time series data of the disease dynamics. A limited volume of synthetic data generated from the COVASIM simulator was used for training the model. Thus we have shown by construction that a prediction of the disease dynamics from effective network properties is possible with sufficient accuracy without incurring the computational cost of performing a full agent based simulation on a complex contact network.

We considered contact restrictions by public health interventions which modify the contact networks in schools, community and work and the corresponding values of complex network measures. The speed-up of predictions by the substitutional model allowed to efficiently sample the space of possible contact restrictions and to quickly predict the corresponding disease dynamics. From a parameter space study an inverse model was set up to derive the manifold in intervention space which is consistent with given external constraints, e.g. a maximum number of critically sick or hospitalized patients. This manifolds marks the weakest contact restrictions compatible with an externally pre-defined maximally acceptable disease dynamics. A discussion of the points of the manifold allows to open a public debate on the least intrusive and most acceptable strategy for infectious disease control.

## II. METHODOLOGY

We use COVASIM to set up realistic contact networks based on a synthetic population which resembles the city of Aschaffenburg, Germany. Its roughly 70,000 inhabitants were represented by 70,000 agents and synthetic contact networks, structured as four subgraphs for households, school, work and communities, were created to reproduce essential macroscopic statistical quantities as implemented in COVASIM. Interaction patterns and rates within and across the subgraphs are based on the synthetic population and modeled through a stochastic process to capture the heterogeneity of disease transmission.

Analysis was conducted in Python version 3.6.15 and Docker. The Docker image includes the required Python packages, COVASIM source code, and any additional data needed for the simulations, and all code used here can be found in: https://github.com/kathlab/covid23.

### A. Creating synthetic data from COVASIM simulation

We use COVASIM to perform a parameter study for varying contact restrictions in schools (s), community (c) and work (w), which implies varying contact networks (Figure 1-I)). 1331 scenarios have been created and the full disease dynamics has been calculated using COVASIM. The household subgraph was excluded from the study as public health interventions in households are considered exceptionally intrusive, their impact is considered less significant [28, 29] and, for practical purposes, to reduce the dimensionality of the parameter space. By concentrating on school, community, and work settings, we captured the aspects of transmission in the public sphere, which is crucial for understanding the effectiveness of most commonly applied intervention strategies [4].

**FIG. 1:**
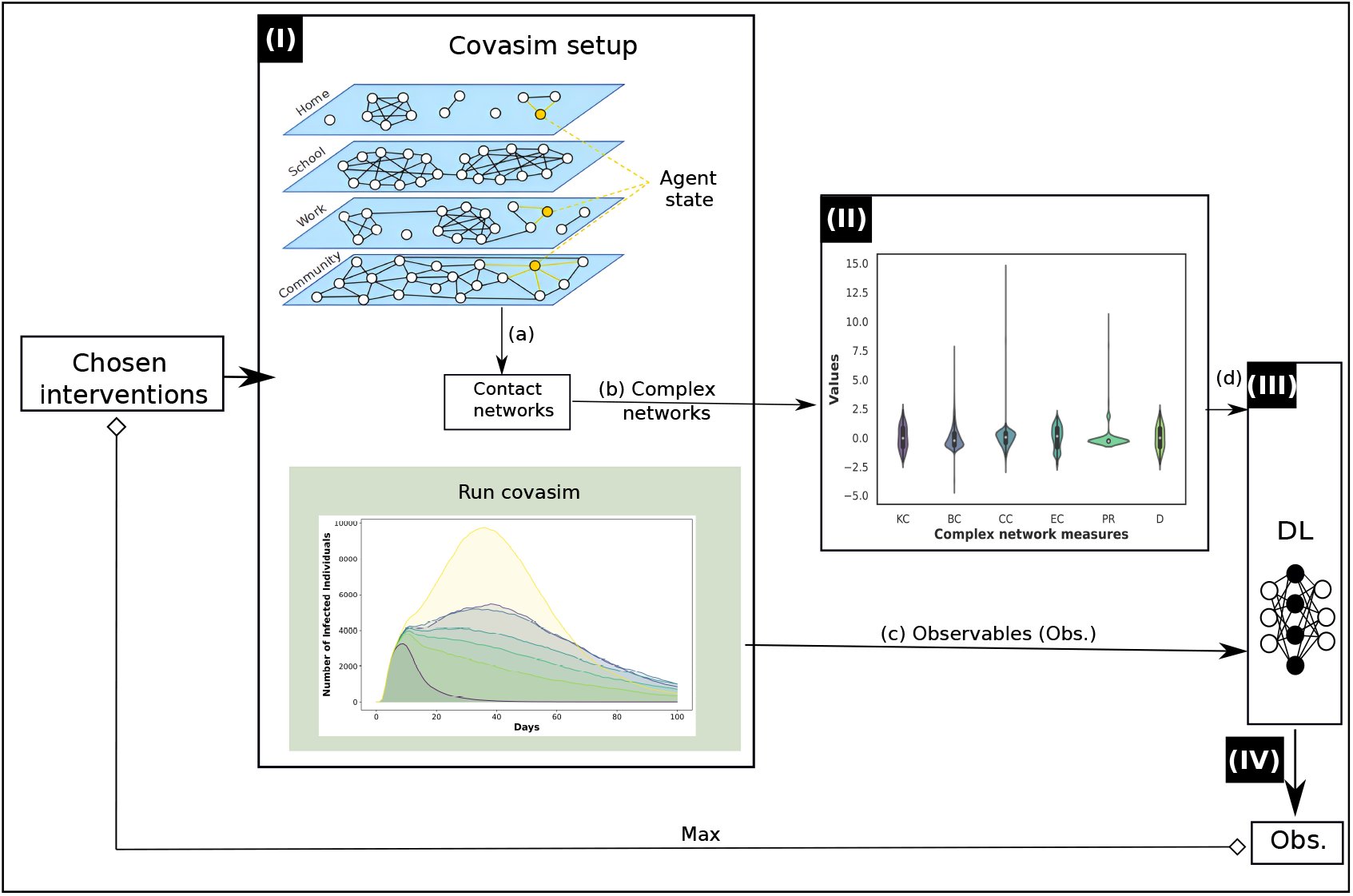
Schematic representation of the methodology using COVASIM simulation. After the chosen interventions, the scenarios are simulated in COVASIM.**(I) Creating synthetic data from COVASIM simulation, described in the subsection II A:** The simulation involves three contact network subgraphs—school, work, and community— jointly represented by a multigraph. Each agent is represented by one node in at least one subgraph (up to maximally one node in each subgraph) and characterized by a multi-valued agent state (in the orange box) as detailed in [4]. Links on the contact network graph represent the possibility for disease transmission between agents which is randomly substantiated through a stochastic process (I.a). We consider 1331 unique variations in the contact network, each differing by the connectivity within the subgraphs. The disease dynamics is represented by time series data of observables (Obs), e.g. the time dependent number of infected agents, and vectorized as I(t). It is calculated using COVASIM for a modeling time of 100 days; exemplary simulation results are illustrated for 7 scenarios (I.b). From the multigraphs, six complex network measures —KC, BC, CC, ECP, R, and D—, which characterize the contact networks, were extracted for dimensionality reduction. The distribution ranges of these measures for the considered networks are depicted in (II). Substitutional ML and DL models (II C) are trained to predict the observables in (II.b) from complex network measure inputs. Finally, the manifold of minimal intervention is recovered as described in II D.

The initial infected population was set at 4.5% of the total population, reflecting early-stage epidemic conditions often observed in urban settings. This percentage was chosen based on epidemiological data suggesting an initial infection seeding rate in similar contexts. For example, the early phases of the COVID-19 pandemic in various European cities saw similar infection rates, as in documented rapid increases in urban areas due to high population density and connectivity. The transmission rate, represented by the parameter *β* = 0.01825, and to differentiate between varying disease severities, we set the relative probabilities of developing severe and critical cases to 0.6558 and 0.9404, respectively. These values were derived from our previous work [31, 32] to calibrate and align with observed COVID-19 transmission dynamics, ensuring that the simulated spread closely mirrors real-world patterns. The simulations of disease dynamics were conducted for 100 days.

We calculated each scenario as an ensemble of five equivalent implementations to account for stochastic variability, enhancing our findings’ statistical robustness; we calculated average outcomes across multiple runs, reducing the impact of random fluctuations and providing more reliable results [4, 33, 34].

### B. Complex network feature extraction for dimensionality reduction

In line with [27], for each of these contact networks the following complex network metrics are extracted; they incorporate essential networks properties and provide dimensionality reduced input features for a machine learning model: Firstly, betweenness centrality (BC), introduced by [35], measures the extent to which a node lies on the shortest paths between other nodes, indicating its role as a bridge or a bottleneck within the network. Closeness centrality (CC), also developed by [36], assesses how close a node is to all other nodes in the network, reflecting the efficiency with which information can spread from that node. Eigenvector centrality (EC), proposed by [37], considers the number of connections a node has and the importance of the nodes it is connected to, providing a measure of a node’s influence within the network. In addition, PageRank (PR), originally devised by [38] for ranking web pages, is employed to evaluate the importance of nodes based on the quantity and quality of incoming connections. Degree (D), discussed by [39], is the simplest measure, representing the number of direct connections a node has, which can indicate its potential for interaction within the network. Finally, the k-core (KC) measure, described in [40, 41], identifies the largest subnetwork in which each node is connected to at least k other nodes, highlighting the network’s cohesiveness. Then, the mean values, which distribution ranges of these measures Figure 1- (II) of the complex network measures were used as the ML features to predict the corresponding I(t).

The ensemble averaged values of the complex network measures, whose distribution ranges can be seen in Figure 1- (II), were used as the ML features to predict the corresponding I(t).

### C. Substitutional ML and DL algorithms

We have employed a multioutput regression methodology which enables the simultaneous prediction of multiple target variables, collectively denoted as I(t). We implemented it for several ML algorithms and compared their performance: the Support Vector Machine (SVM) algorithm, as proposed by [42]; the Random Forest (RF) algorithm [43]; and the scalable tree boosting algorithm (Xgboost) [44]. We employ grid search for hyperparameter tuning with mean R2 score as the optimizing criterion. [45–50]. The set of hyperparameters and range of values considered in the grid search is shown in Table I. The synthetic dataset was split into disjunct training (75%) and test (25%) subsets and training was performed w.r.t. the target variables I(t).

**TABLE I:** Hyperparameters for different machine learning model.

| Model | Hyperparameter | Values | Description |
| --- | --- | --- | --- |
| SVM | estimator C | 0.1, 1, 10 | Regularization parameter |
|  | kernel | linear, rbf | Kernel function |
| RF | maximum depth | 10, 20, 30, 40, 50 | Maximum depth of each tree |
|  | maximum features | 2, 3, 4 | Maximum number of features for the best split |
|  | number estimators | 100, 200, 300 | Number of trees in the forest |
| MLP | hidden layer sizes | (100,), (100, 50, 100) | Number of neurons in the hidden layers |
|  | alpha | 0.0001, 0.001, 0.01 | L2 penalty (regularization term) |
|  | solver | adam, SGD, lbfgs | The solver for weight optimization |
| XGBoost | n_estimators | 100, 200, 300 | Number of boosting rounds |
|  | learning rate | 0.01, 0.1, 0.2 | Step size shrinkage |
|  | max depth | 3, 5, 7 | Maximum depth of a tree |

Model performance was measured using the conventional R-squared metrics[51–53], to assess the goodness-of-fit of our predictive models. The R2 score measures the proportion of the variance in the dependent variables that our models can elucidate.

Additionally, a previous split of 25% is made on the original dataset, reserved for final testing after the model is trained on the 10-fold cross-validation. This technique evaluates model performance while minimizing overfitting and ensuring generalization to new data Moreover, we implemented a fully connected neural network as deep learning model (DL). To efficiently tune the hyperparameters of this model, we used the random search optimization algorithm, which involves randomly sampling hyperparameters from a predefined range and evaluating the model’s performance for each set [54]. This method effectively identifies satisfactory hyperparameter configurations without requiring exhaustive searches across the entire hyperparameter space [55]. The high dimensionality and complex interplay of hyperparameters in deep learning render traditional grid search impractical [56, 57].

Additionally, we applied dropout and L2 regularization techniques to mitigate overfitting. Dropout involves temporarily deactivating a random subset of neurons during each training iteration, preventing the model from becoming overly dependent on specific neurons and thereby enhancing generalization [58]. L2 regularization, also known as weight decay, was applied to the weights of the neural network layers. Additionally, we applied dropout and L2 regularization techniques to mitigate overfitting. Dropout involves temporarily deactivating a random subset of neurons during each training iteration, preventing the model from becoming overly dependent on specific neurons and thereby enhancing generalization [58]. L2 regularization was applied to penalize larger weight values and to reduce the risk of overfitting [59]. The architecture and random search hyperparameters for the DL algorithm are summarized in Table II.

**TABLE II:** DL algorithm architecture and hyperparamter optimization.

| Layer | Hyperparameter | Value Range |
| --- | --- | --- |
| Dense | Units | 8, 12, 16, . . . , 60, 64 |
|  | Activation function | relu, tanh, sigmoid |
| Dense | Units | 16, 20, 24, . . . , 60, 64 |
|  | Activation function | relu, tanh, sigmoid |
| Dropout | Rate | 0.001, 0.02, 0.03, . . . , 0.59, 0.6 |
| Dense | Units | Number of time series |

We consistently applied a uniform data sampling strategy across all machine learning and deep learning algorithms, using a 10-fold cross-validation approach with shuffling; k=10 is a common value for this method [60– 64]. This technique involves partitioning the dataset into ten equitable folds, ensuring each fold represents a fair portion of the data. Random shuffling ensures that each batch contains a different mix of data points in every iteration. Shuffling the data before partitioning reduces the risk of any systematic patterns in the dataset influencing the performance of the model. This process is suitable to reduce a possible bias, enhance the robustness of the trained models and to improve generalization capabilities to unseen data[65–67].

Furthermore, per established preprocessing best practices, our pipeline incorporates the application of a standard scaler to normalize the features before training our multioutput model. Following the approach outlined in [68, 69], we employed this scaler to standardize the features by centering them through the removal of the mean and subsequently scaling to attain unit variance. This normalization procedure proves pivotal, especially for algorithms that hinge on the assumption of feature homogeneity. Both the input features (X) and the corresponding multioutput target variables (y) are subjected to this scaling protocol, which is recognized for its capacity to enhance the suitability of the data for a diverse array of machine learning algorithms and optimization methodologies. This uniformity in scaling fosters heightened model convergence and stability and culminates in demonstrably enhanced performance.

The SHapley Additive Explanations (SHAP) values were calculated to assess the predictive contribution of individual variables, the complex network metrics described in subsection II A. The SHAP value methodology, rooted in cooperative game theory and popularized by [70], allows to conclude on the relative contributions of each input feature (here: the complex network measures) to the model prediction (here: prediction variables I(t)). For details see III A.

### D. Assessing the manifold of minimal intervention

For illustration, we consider a use case where disease control should be done with minimal restrictions but within a set of constraints set by the health system. Such constraints on the maximally acceptable number of infected, hospitalized or critically sick patients are given externally. For instance, the number of available beds in intensive care units can be a limiting factor which requires to keep the number of critically sick patients below that threshold. That goal, however, may be equivalently reachable by different contact network restrictions applied to one or several subgraphs of the contact network. Therefore, we consider the manifold of equivalent and minimal interventions which all imply a disease dynamics compatible with the set constraints.

Motivated by this use case we restrict our target variables (time series data on disease dynamics) to the maximum value of critically sick patients at any point in prediction time to facilitate the assessment of optimal interventions. We consider the three dimensional parameter space defined by the strength of the restrictions imposed on the school, community or work subgraphs and, as a single target value, the maximum number of the critically sick. The trained deep learning model was used to refine the parameter space discretization and to extend the synthetic dataset. Cubic splines [71] were used to finally interpolate the manifold [72] between available data points.

Figure 1 depicts our methodology summary scheme.

## III. RESULTS

### A. Machine learning and deep learning results

Figure 2-(a) depicts the results of ML and DL algorithms. By construction we verified that the time series of observables can be predicted directly from complex network measures as input features, bypassing a full agent based simulation on a contact network, with reasonable accuracy; all algorithms obtained R2 test higher than 85%. The DL algorithm achieved the best performance, with a mean train R2 of 0.9587 ± 0.0076 and a test R2 of 0.9605.

**FIG. 2:**
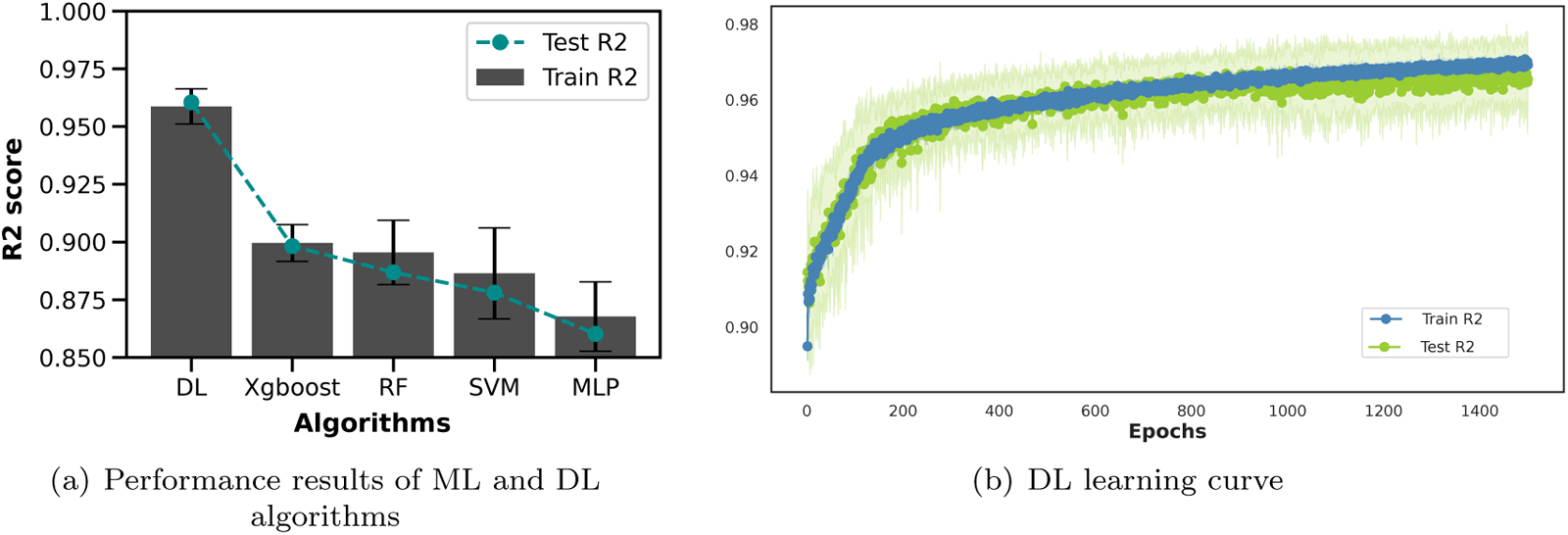
Performance Comparison of ML and DL Algorithms. (a) R2-performance for ML and DL algorithms. The grey bar plot represents each algorithm’s training performance, including error bars. The blue curve indicates the R2 performance for the test data. (b) The learning curve for the optimized DL model the plot displays the mean R2 scores for both train (blue) and test (green) sets, shaded regions the standard deviation.

Figure 2-(b) illustrates the learning curves over epochs for the DL model. An epoch represents one complete pass through the entire training dataset. The curve shows the development of the model performance with ongoing training, i.e. updates to the trainable weights over multiple iterations. Both the training and test performances of the DL models increase until convergence, indicating a stable and well-generalized model.

The convergence of learning curves in DL models indicates that this model achieved stable performance, which is crucial for reliable predictions and generalization to new data.

Further, we also fit the flattened predicted time series found (ŷ) and compared it to the flattened original data (y), as shown in Figure 3. It can be seen that the cosine similarity between the predicted values and the original data is higher than 99%, demonstrating the high predictive accuracy of our DL model. We conclude that complex network measures are a suitable candidate for a reduced representation of contact networks which still maintains predictive power.

**FIG. 3:**
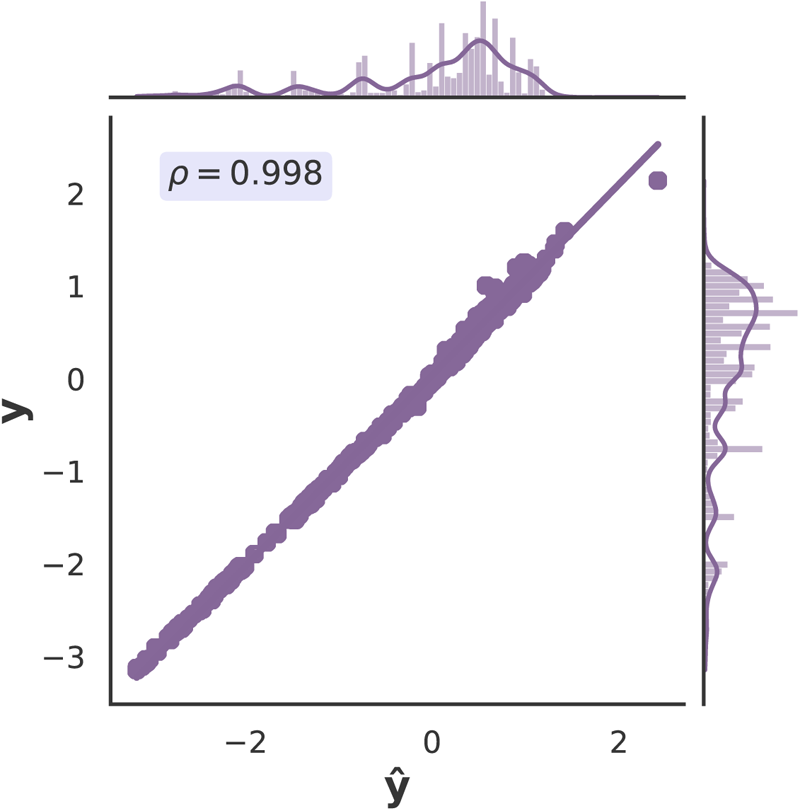
Comparison of flattened predicted time series and flattened original data for DL model. The figure presents the flattened predicted time series (ŷ) compared to the flattened original data (y) across various algorithms. Both ŷ and y were normalized using standard scalers before the comparison. The cosine similarity (*ρ*) was calculated for each comparison to quantify the similarity between the predicted and original series.

Since the DL model obtained the best performance, the SHAP value analysis is applied to this model, depicted in Figure 4, revealing the relative importance of various complex network metrics in predicting the number of infected individuals over time, I(t). The bar plot (Figure 4-a) indicates that PR has the highest impact on the model’s output, suggesting that the influence of a node, considering the importance of its neighbors, plays a crucial role in understanding the spread of infection. Following PR, CC is also highly influential, highlighting the importance of nodes’ accessibility to others within the network.

**FIG. 4:**
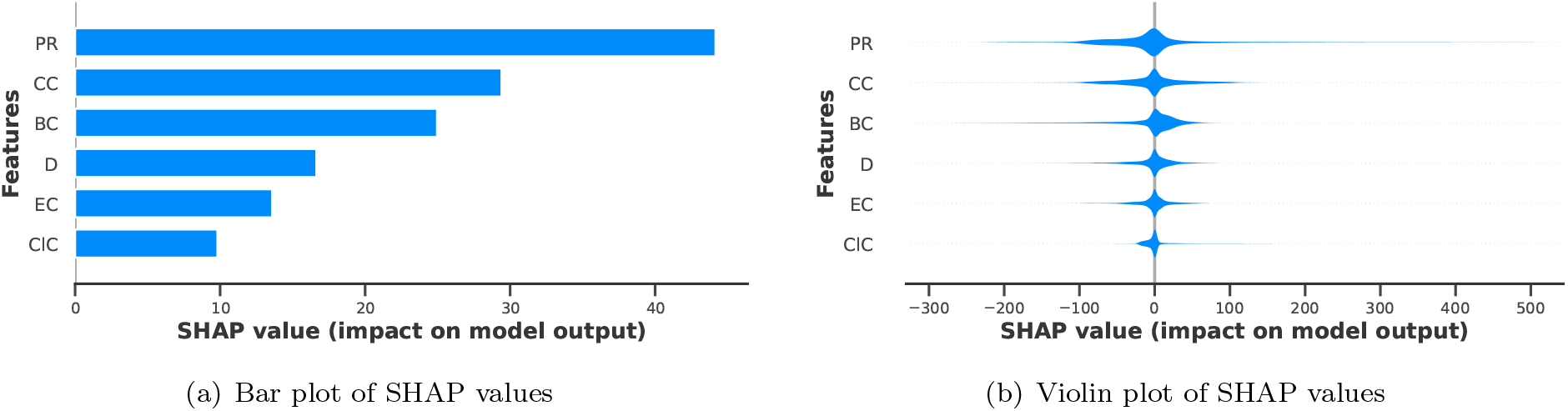
SHAP value analysis for the deep learning model. (a) The bar plot shows the mean SHAP values for each complex network metric, indicating their average impact on model predictions. PR has the highest impact, followed by CC, BC, D, EC, and KC. (b) The violin plot shows the distribution of SHAP values for each feature, illustrating the variability in their impacts across different scenarios. Broader distributions for PR and CC suggest higher variability, while narrower distributions for BC, D, EC, and KC indicate more consistent impacts.

The violin plot (Figure 4-b) complements the bar plot by showing each feature’s distribution of SHAP values. PR and CC have broader distributions, indicating varying impacts across different scenarios. This variability suggests that while these features are generally significant, their influence can differ significantly depending on the specific network structure and intervention scenario. The distributions for BC, D, EC, and KC are narrower, indicating more consistent impacts across different scenarios.

Further, the DL model is applied to time series for critical and severe patients and the infected patients’ time series. Table III depicts the results of DL to the all I(t) curves in which similar results can be obtained for the infected patients. This consistency suggests that the DL models achieve stable performance across different patient categories, which is crucial for reliable predictions and generalization to new data. Further, the severe and critical time series learning curves can be seen in Appendix B, whose curves depicted the convergence and stability of the DL model for different curves.

**TABLE III:** DL performance to different I(t) curves.

| $I(t)$ | Train Mean R2 | Test R2 |
| --- | --- | --- |
| Infected | $0.9587 \pm 0.0076$ | 0.9605 |
| Severe | $0.9738 \pm 0.0037$ | 0.9824 |
| Critical | $0.9841 \pm 0.0027$ | 0.9820 |

### B. Assessing the manifold of minimal intervention

As described in subsection II D, using spline interpolation in the intervention space (S,c,w) with the maximum number of critically sick as the target variable, we mapped out the manifold of minimal intervention. Figure 5 displays three cross sections through 3D parameter space. It illustrates the manifold for different public health interventions which restrict contacts in any combination of the dimensions community, work, and school and allows to read off the effectiveness of actions. Various intervention strategies can be discussed w.r.t. their impact on the maximum number of critical cases: The subfigures (A), (B), and (C) depict the combined impact and the interplay between restrictions in community and work work (with school set to 0), community vs. school (with work set to 0), and school vs. work (with community set to 0), respectively. The color gradients represent the maximum critical cases observed under each intervention scenario, highlighting the regions that result in the lowest and highest number of critical patients. This visualization aids in identifying equivalent interventions which are consistent with a pre-defined maximal number of critical cases. This enables a broader discussion on the optimal public health response in a society. A full 3D visualization of the considered parameter space is depicted in Appendix C.

## IV. DISCUSSION

In this study we, firstly, investigated the research question whether a full simulation of disease dynamics using an agent based simulator is necessary to predict the time series of essential observables like the number of infected, hospitalized or critically sick patients.

By applying machine learning to a set of dimensionality reduced features of the contact network, defined by complex network measures, we observed that models with equivalent predictive power can be constructed. A DL approach achieved the highest performance, with an R2-adjusted value exceeding 95%. This high performance signifies the model’s efficiency in predicting the time series of infected, severe, and critical cases within a sufficient approximation. This result confirmed our hypothesis that the topology of the contact network could effectively characterize the impact of the network on the disease dynamics. Therefore, we successfully abstracted from the full description based on an adjacency matrix of the contact network to effective network metrics; this allows -at least partially- to circumvent the need for computationally expensive agent-based simulations.

The SHAP value results are useful to understand the relative relevance of the applied complex network measures. It demonstrates that centrality measures, mainly PR and CC, are leading in capturing the nuances of disease transmission within the network. PR is vital as it accounts for both the quantity and significance of a node’s connections, aiding in identifying influential nodes [73–75]. CC measures how quickly an infection can spread from one node to all others, identifying nodes crucial for rapid dissemination [76, 77]. Our findings align with the study [78], emphasizing the predictive power of combined centrality measures. The study demonstrated that integrating normalized spectral centralities like PR with measures sensitive to graph edges, such as CC, can yield rather high predictive accuracy (R2 scores of 0.91 or higher) across various graph structures and epidemic parameters. This reinforces the notion that PR’s consideration of both the quantity and quality of connections, coupled with CC’s ability to measure rapid dissemination potential, makes them highly effective for identifying influential nodes and optimizing intervention strategies. Our use of these centralities indicates their applicability and robustness in network epidemiology.

Further, BC and D also show substantial contributions, emphasizing the roles of nodes in controlling information flow [35, 79] and the number of direct connections, respectively [80]. EC and KC have slightly lower SHAP values but contribute significantly to the model predictions. EC reflects the influence of nodes based on the quality of their connections [37]. At the same time, KC indicates the core-periphery structure of the network, both providing valuable insights into the network’s robustness and connectivity [40].

These findings validate our DL model’s robustness and ability to effectively leverage network-based features to predict disease dynamics. This enhances our understanding of intervention strategies and their potential impacts on public health, supporting the development of more effective containment and mitigation measures.

A further research question directs towards a better understanding of equivalent public health interventions obtained through contact restrictions in various contexts of society. A critical aspect of our study is to identify the manifold of minimal intervention for a given set of external constraints, illustrated by a maximal number of critically sick patients. Through extensive parameter space sampling and analysis, we mapped out the relative impact of contact restrictions in schools, workplaces and community on disease dynamics (in Figure 5). This provides a route towards improved tools for policymakers and the public alike to decide on the best strategies based on their specific public health constraints but also on an appropriate sharing of burden. The results indicate that interventions in the community and work layers are equally effective for controlling the pandemic, as demonstrated by the diagonal line representing balanced interventions between these two layers. Specifically, within the community vs. work intervention space (in Figure 5), a coordinated contact reduction in both areas yields the lowest number of critical cases. Community interventions are more effective in reducing critical cases when comparing community and school interventions (in Figure 5). When resources or compliance levels are limited, public health strategies should prioritize community-based measures over school closures. Conversely, the comparison between school and work interventions (in Figure 5) reveals that workplace interventions generate more impact in controlling the pandemic than school-based measures. This may raise questions on the relevance, necessity and appropriatness of large scale school closures to mitigate the spread of the virus. Figure 5 supports the conclusion that both community and work interventions are of comparable impact. This enables mutual trade-offs and allows for a societal debate on which combination of contact restrictions, chosen from equally effective ones, may be most appropriate for disease control in a pandemic situation.

**FIG. 5:**
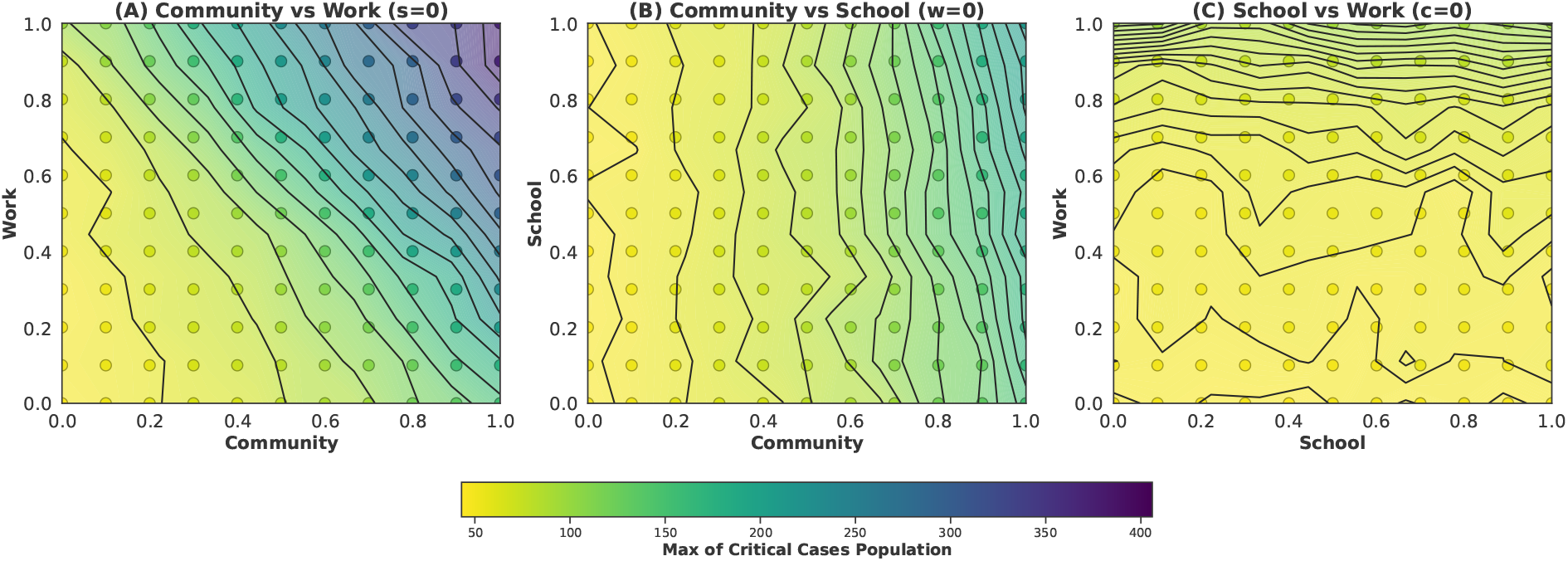
Cross section representation of varying contact restriction parameters. The color coding indicates the maximum number of critical cases, with lighter colors representing lower numbers and darker colors indicating higher numbers of critical cases. Lines of constant values of critical cases are printed to illustrate dependencies. Parameters describe the restrictions on the corresponding subgraphs. (A), (B), and (C) shows pairwise dependencies, keeping the third parameter fixed.

In summary, our findings on the relative importance of contact restricting interventions indicate that among school, community, and work settings, community restrictions have the most significant impact on controlling the disease dynamics. This aligns with the general understanding that casual and frequent interactions in community settings contribute substantially to disease transmission [81, 82]. The analysis also highlights that contact restrictions at workplaces generate a higher impact than school closures. We attribute this, among other aspects, to the high density and regularity of contacts in workplace environments.

However, despite these promising results, several limitations must be acknowledged. First, the chosen approach was applied to a very limited parameter space, only addressing changes to contact networks described by three parameters. We maintained constant all other parameters available in COVASIM, in particular medical parameters, such as transmission and recovery rates, across all simulations. These parameters can vary significantly depending on local health conditions, population behavior, and emerging virus variants. Second, our model does not account for the impact of other types of interventions like vaccinations, active contact tracing or others that have become crucial in managing pandemics. The exclusion of household contacts might oversimplify the contact dynamics, although we focused on public and communal settings to capture broader transmission patterns. Lastly, our approach relies heavily on the detection of network modifications stemming from contact restrictions by established complex network measures. While this is the case in the currently depicted contact restriction scenarios, it may happen that, in reality, different modifications of contact networks occur which may not be equally well resolved by complex network measures. A test of the approach with real network data is, unfortunately, beyond the scope of this study but could provide further evidence on the reliability of the chosen route. We mention that we have not exhausted the set of possible complex network measures and that further measures could be added to the set of input features if required.

## V. CONCLUSION

The COVID-19 pandemic has highlighted the need for accurate and efficient models to predict disease dynamics and guide public health interventions. This study utilized the COVASIM agent-based model to simulate various scenarios across different social settings—specifically focusing on school, community, and workplace related contact networks. By extracting complex network measures from these simulations and applying deep learning algorithms, we were able to predict key epidemiological outcomes, including the number of infected, severe, and critical cases, with a high degree of R2 values exceeding 95%.

Our approach demonstrated robust predictive capabilities and provided a framework for identifying optimal intervention strategies. We observed that community and workplace interventions are critical in minimizing the impact of the pandemic, underscoring the importance of targeted public health strategies in these areas. The integration of network analytics with deep learning offers significant advantages in epidemic modeling, including reduced computational costs and enhanced decisionmaking efficiency.

While our model successfully abstracted the complex dynamics of disease transmission, it’s important to acknowledge certain limitations. These include the assumption of constant medical parameters, excluding vaccination effects and others. This underscores the need for ongoing research to incorporate dynamic parameters and a broader range of contact layers, which will further improve the model’s applicability and robustness.

In conclusion, this study provides a novel and effective framework for planning most adequate contact restrictions to control infectious disease outbreaks. By leveraging deep learning and network measures, our approach can simplify parameter space searches and offers valuable insights for public health decision-making.

## DATA AVAILABILITY STATEMENT

The data that support the findings of this study are available from the corresponding author upon reasonable request.

## Appendix A: Intervention space

Figure 6 illustrates the intervention space used for the COVASIM simulation.

**FIG. 6:**
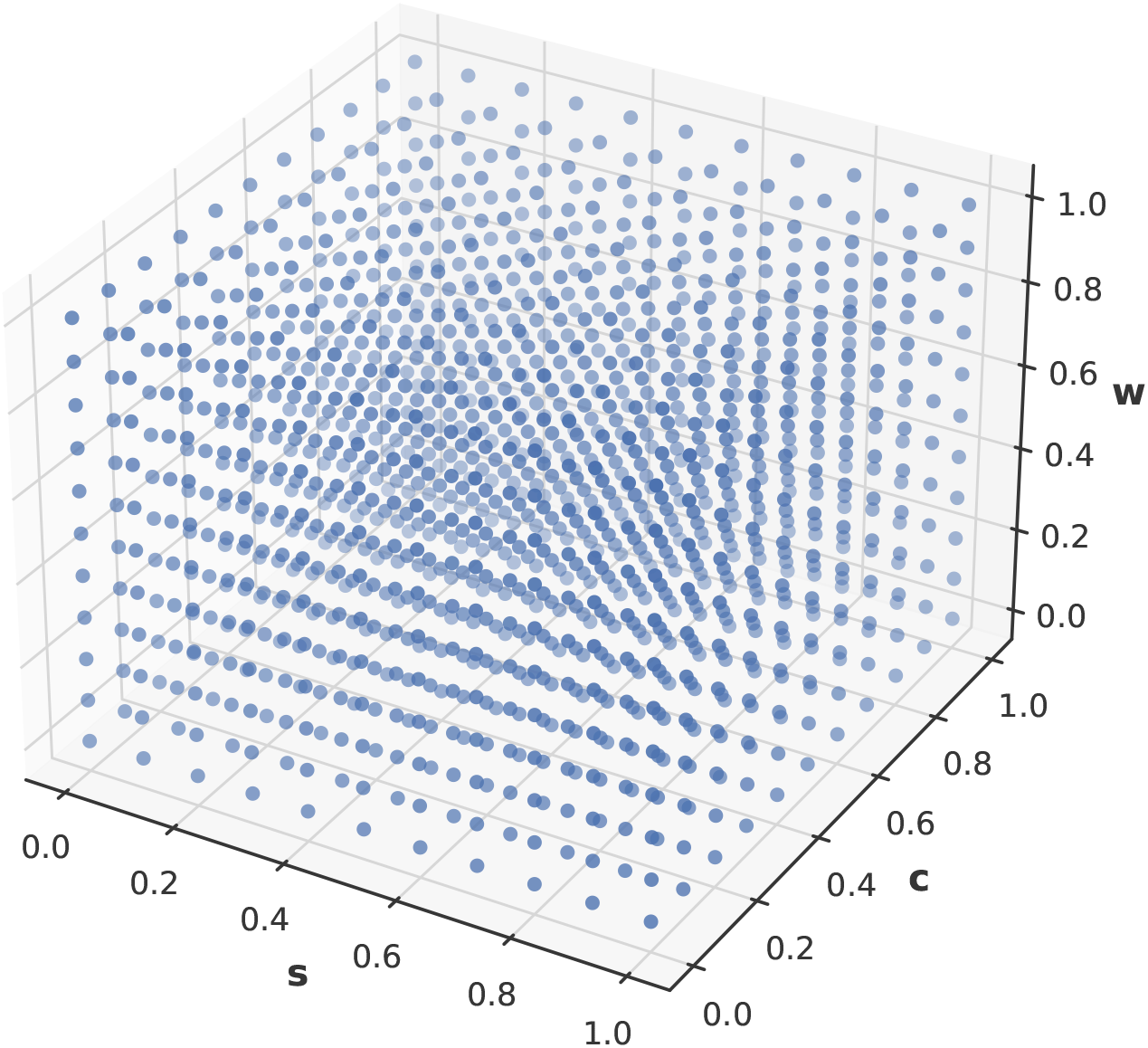
A 3D intervention space. The intervention space for the COVASIM simulation with 1331 different scenarios used in the present work in each coordinate represents a layer: school (s), community (c), and work (w).

## Appendix B: Learning curves for different I(t)

The following figures illustrate the learning curves for the DL model applied to the time series of severe and critical patients, alongside the infected patients’ time series. Each plot depicts the mean R2 scores for both train and test sets, with shaded regions representing the standard deviation. These curves highlight the model’s performance stability across different patient categories, reinforcing the similar results obtained.

**FIG. 7:**
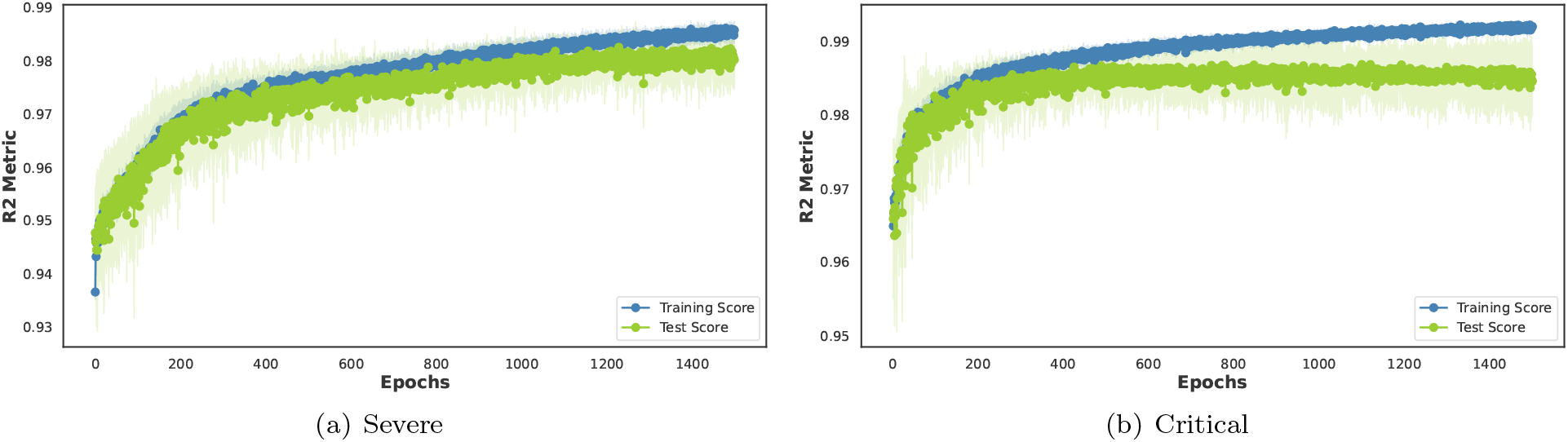
Learning curves for severe (a) and critical (b) patient time series using the R2 score. The plots display the mean R2 scores for both train and test sets (in blue and green, respectively), with shaded regions indicating the standard deviation.

These learning curves provide a visual representation of the model’s training and validation process, demonstrating how the performance stabilizes over time. The convergence to stable R2 scores in both training and test sets across different patient categories emphasizes the robustness and reliability of the DL model in making accurate predictions.

## Appendix C: Assessing the surface with optimum interventions with DL generated scenarios

Figure 8 shows the interpolation of intervention effects on the maximum number of critical cases, generated using our deep learning model. The curve maintains the same overall pattern observed in Figure 8, but with increased detail due to the higher number of data points from the deep learning model. This detailed interpolation allows for a more nuanced understanding of the optimal intervention strategies, providing clearer insights into the relationships between community, work, and school interventions and their impact on critical cases.

**FIG. 8:**
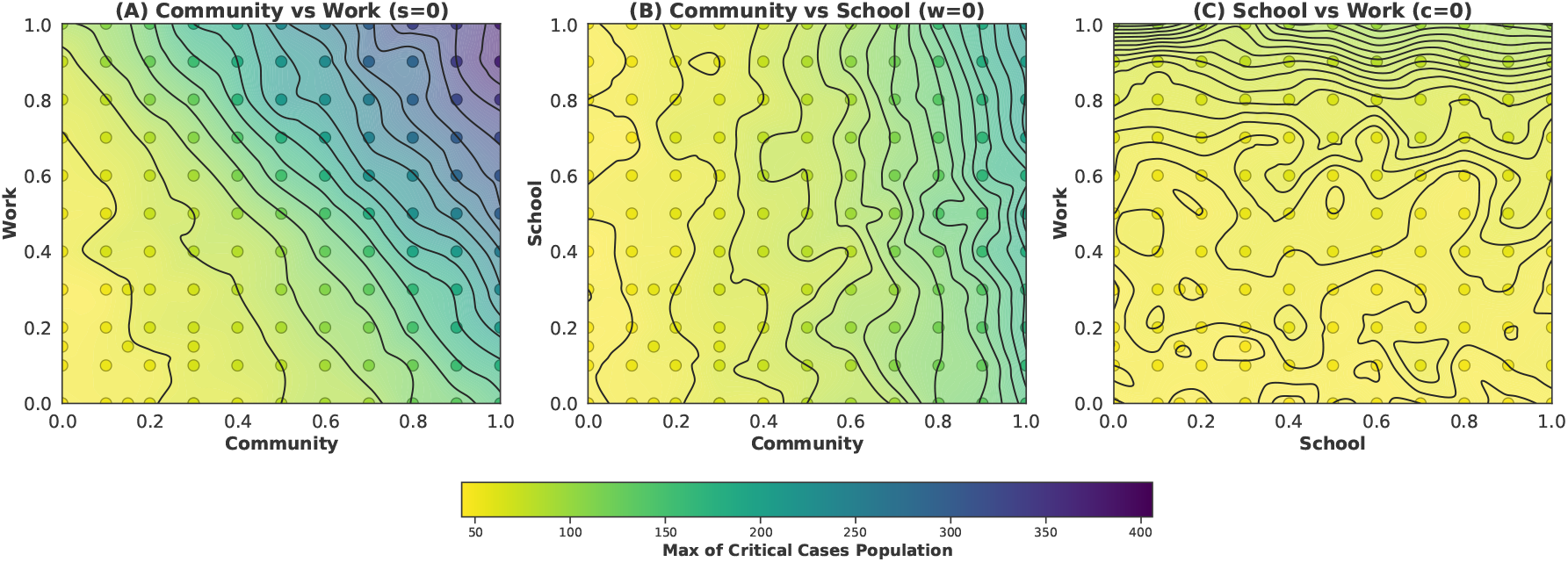
Interpolation of intervention effects on the maximum number of critical cases using DL-generated scenarios. The increased number of data points results in a more detailed interpolation, enhancing the understanding of optimal intervention strategies.

